# Developing multifactorial dementia prediction models using clinical variables from cohorts in the US and Australia

**DOI:** 10.1101/2024.03.12.24304189

**Authors:** Caitlin A. Finney, David A. Brown, Artur Shvetcov, the Alzheimer’s Disease Neuroimaging Initiative, the Australian Imaging Biomarkers and Lifestyle Flagship Study of Ageing

## Abstract

**INTRODUCTION:** Existing dementia prediction models using non-neuroimaging clinical measures have been limited in their ability to identify disease. This study used machine learning to re-examine the diagnostic potential of clinical measures for dementia.

**METHODS:** Data was sourced from the Australian Imaging, Biomarkers, and Lifestyle Flagship Study of Ageing (AIBL) and the Alzheimer’s Disease Neuroimaging Initiative (ADNI). Clinical variables included 21 measures across medical history, hematological and other blood tests, and APOE genotype. Tree-based machine learning algorithms and artificial neural networks were used.

**RESULTS:** APOE genotype was the best predictor of dementia cases and healthy controls. Our results, however, demonstrated that there are limitations when using publicly accessible cohort data that may limit the generalizability and interpretability of such predictive models.

**DISCUSSION:** Future research should examine the use of routine APOE genetic testing for dementia diagnostics. It should also focus on clearly unifying data across clinical cohorts.

## 1. BACKGROUND

More than 55 million people worldwide have dementia, a number that is rapidly rising by an additional 10 million cases yearly [1]. Dementia is the leading cause of disability among older adults over the age of 65 and costs the global economy more than $1.3 trillion USD annually [1]. The timely diagnosis of dementia is essential to ensure that patients and their families can access early support and interventions, thereby improving quality of life, prolonging independence, and reducing the healthcare burden [2]. Improving dementia diagnostic capabilities and rates, therefore, is an urgent priority [1, 3].

The recent advent of increased computational power and of ‘big data’ from large clinical cohorts has led to an increase in machine learning models for developing diagnostic tools to identify dementia. Most current models have relied on neuroimaging from magnetic resonance imaging (MRI) and positron emission tomography (PET) scans [4, 5]. Despite showing a high level of performance and predictive power, practicality constraints limit the day-to-day clinical usefulness of such models. Importantly, neuroimaging is difficult for many patients to access to due factors including health insurance status, out-of-pocket costs that limit their affordability, and if the patient lives rurally or in a major metropolitan area [2, 6, 7]. Additionally, waitlists for neuroimaging can be upwards of several months and require specialist services for processing and interpretation, leading to delays in dementia diagnosis [7]. These issues were further highlighted by the recent Alzheimer’s Association Primary Care Physician Dementia Care Training Survey that found more than half of primary care physicians feel they do not have the local specialist resources to meet patient demand [8]. In line with these practical constraints, machine learning models based on diagnostic imaging have had limited utility in real world clinical applications [6].

Primary care physicians are essential for patient triage, diagnosis, and management [2]. Therefore, from a practical perspective, machine learning-based dementia diagnostic models should focus on the predictive power of easy-to-obtain clinical measures. Indeed, previous studies have used machine learning to look at the diagnostic utility of routine clinical measures including, for example, the Cardiovascular Risk Factors, Aging, and Dementia (CAIDE) [9], Study on Aging, Cognition and Dementia (AgeCoDe) [10], Australian National university Alzheimer’s Disease Risk Index (ANU-ADRI) [11], Rapid Assessment of Dementia Risk (RADaR) for older adults [12], and Brief Dementia Screening Indictor (BDSI) [13]. There are limitations, however, to these models. First, some models used clinical variables that were used to define a dementia patient relative to a healthy control, including the Mini-Mental State Exam (MMSE) and Clinical Dementia Rating (CDR) scores [14, 15]. In machine learning models, this constitutes “data leakage” where an experimental group label (e.g. dementia defined by MMSE <24) is also included as a characteristic (MMSE scores per person) in the model. This leads to an artificially improvement in model performance while significantly limiting generalizability and interpretability. Second, although these models report a high specificity (identification of true healthy controls) and negative predictive value (NPV; ratio of true healthy controls to all healthy controls identified), they report very low sensitivity (identification of true dementia patients) and positive predictive value (PPV; ratio of true dementia patients to all dementia patients identified [9, 14, 16, 17]. Reported sensitivities and PPVs ranged from 0.1-0.47, indicating that models using clinical measures can readily identify a healthy person but are unable to identify someone with dementia. In direct support of this, a recent study confirmed that these existing dementia prediction models missed 84-91% of patients with incident dementia therefore demonstrating little, if any, clinical utility for dementia diagnostics [18]. This underlies the need for the development of sensitive prediction models that identify patients with dementia.

In this context, using two large cohorts from Australia (the Australian Imaging Biomarkers and Lifestyle Flagship Study of Ageing; AIBL) and the US (Alzheimer’s Disease Neuroimaging Initiative; ADNI), the primary aim of this study was to use several different machine learning models to re-examine the diagnostic potential of easy-to-obtain clinical measures for dementia in older adults over age 65.

## 2. MATERIAL AND METHODS

### 2.1. Data and participants

The data used in the present study was obtained from two publicly available databases: ADNI and AIBL (https://ida.loni.usc.edu/) and the data was downloaded on October 10, 2023. Participants included those enrolled in ADNI and AIBL who were identified as either a healthy control or diagnosed dementia patient (probable Alzheimer’s disease (AD)) at their baseline visit and assessments. ADNI participant characteristics have been described elsewhere [19]. In brief, those with a dementia diagnosis were identified as having subjective memory complaints, an MMSE range of 20-26, and CDR of >0.5 [19]. AIBL participant characteristics have also been described elsewhere [20]. Here, participants were identified as having dementia (probable AD) as defined by National Institute of Neurological and Communicative Diseases and Stroke/Alzheimer’s Disease and Related Disorders Association (NINCDS-ADRDA) criteria [21]. AIBL study methodology has been reported previously [20]. Unlike ADNI, AIBL included participants who expressed a concern about their memory function or had memory complaints in response to being asked “Do you have difficulties with your memory” in the healthy control group. This resulted in approximately 50% of the healthy control group made up of those who had subjective concerns about their memory [20]. Participants in both the ADNI and AIBL cohorts that were diagnosed with mild cognitive impairment (MCI) were excluded from the current study as MCI doesn’t necessarily progress to dementia.

Prior to undertaking any analysis, we limited both datasets to participants over age 65 to capture older adults who were more likely to have age-associated dementia, rather than early onset. We identified that there was a statistical difference in the ages between the ADNI and AIBL cohorts with average participant ages of 76 and 74 years, respectively (Wilcoxon test; W = 346983, *p* = 3.17e-11). To remove this age bias between the cohorts, we created a random sample of 150 ADNI participants within the age range of 80-96 and removed them to eliminate this statistical difference. After doing this, the average age was an equal 74 across both the ADNI and AIBL cohorts. Table 1 shows the demographic and clinical characteristics of the participants included in this study after controlling for age.

**Table 1.** Demographic and clinical characteristics of the ADNI and AIBL cohorts included in the current study.

|  |  | ADNI |  | AIBL |  |
| --- | --- | --- | --- | --- | --- |
|  |  | Dementia | Healthy control | Dementia | Healthy control |
| Total N |  | 476 | 292 | 107 | 524 |
| Sex (M:F) |  | 257:185<br>34 (Unk) | 133:153<br>6 (Unk) | 48:59 | 223:301 |
| Age |  | 75 (8) | 74 (6) | 77 (9) | 73 (8) |
| APOE Genotype | e2,e2 | 0 | 3 | 0 | 2 |
|  | e2,e3 | 12 | 45 | 7 | 75 |
|  | e2,e4 | 12 | 4 | 2 | 10 |
|  | e3,e3 | 128 | 173 | 23 | 307 |
|  | e3,e4 | 242 | 60 | 55 | 113 |
|  | e4,e4 | 82 | 7 | 20 | 17 |
| Medical History (Yes:No) | Psychiatric | 204:272 | 75:217 | 33:74 | 100:424 |
|  | Neurologic | 148:328 | 85:207 | 27:80 | 49:475 |
|  | Cardiovascular | 336:140 | 203:89 | 44:63 | 226:298 |
|  | Hepatic | 16:460 | 13:279 | 3:104 | 20:504 |
|  | Musculoskeletal | 316:160 | 208:84 | 42:65 | 283:241 |
|  | Endocrine-Metabolic | 208:268 | 121:171 | 16:91 | 101:423 |
|  | Gastrointestinal | 203:273 | 144:148 | 27:80 | 145:379 |
|  | Renal-Genitourinary | 227:249 | 121:171 | 0:107 | 30:494 |
|  | Malignancy | 106:370 | 72:220 | 16:91 | 90:434 |
| Hematological Tests | Red blood cell count (RBC) | 4.7 (0.5) | 4.6 (0.6) | 4.5 (0.4) | 4.5 (0.5) |
|  | White blood cell count (WBC) | 6.3 (1.9) | 6.3 (2.1) | 5.8 (1.7) | 5.6 (1.8) |
|  | Platelets | 230.0<br>(81.0) | 241.5<br>(76.0) | 237.0<br>(62.5) | 216.0<br>(67.0) |
|  | Hemoglobin | 13.95<br>(1.35) | 13.80<br>(1.60) | 13.8 (1.25) | 14.0<br>(1.40) |
|  | Mean corpuscular hemoglobin (MCH) | 30.0 (2.0) | 30.0<br>(2.0) | 31.2 (1.6) | 31.3<br>(1.8) |
|  | Mean corpuscular hemoglobin concentration (MCHC) | 33.0 (1.0) | 33.0<br>(1.0) | 34.0 (0.8) | 33.9<br>(0.8) |
| Other Blood Tests | Vitamin B12 | 449.0<br>(271.0) | 456.5<br>(331.8) | 448.0<br>(244.6) | 428.3<br>(237.9) |
|  | Serum glucose | 96.0 (17.0) | 96.0<br>(15.0) | 90.1 (11.7) | 90.1<br>(12.6) |
|  | Cholesterol (high performance) | 194.0<br>(55.2) | 193.5<br>(48.5) | 216.5<br>(56.1) | 204.9<br>(50.3) |
|  | Urea nitrogen | 18.0 (7.0) | 18.0<br>(6.0) | 34.8 (9.3) | 36.6<br>(12.0) |
|  | Creatinine | 1.0 (0.3) | 0.9 (0.3) | 0.8 (0.2) | 0.8 (0.2) |

For continuous features (age and tests), the median and interquartile range (in brackets) are shown.

### 2.2. Feature selection (predictors)

We first ensured that the features (or predictive variables) used in our models matched across the ADNI and AIBL cohorts. Common features across both cohorts included apolipoprotein E (APOE) genotype, nine medical history questions, and test results from six hematological and five blood tests. These features along with the median and interquartile range (IQR) across participants and cohorts are shown in Table 1. Although both cohorts had data for MMSE and CDR, these were not used as features due to the potential for data leakage and artificial inflation of model performance, as discussed in the Background. We also had to remove two variables from our analyses: thyroid stimulating hormone test (AXT117) and a health history of smoking (MH16SMOK). Both variables were removed due to too many participants missing data for these. Thyroid stimulating hormone test results were missing for 45% of the ADNI cohort and smoking history was missing for 42% of the AIBL cohort. For the remaining variables, we imputed the missing values where required using the group median value. Of note, neither ADNI nor AIBL specify the units of measurement for the hematological and other blood tests. It is not clear, therefore, if the units of measurement are the same across them. We therefore treat these variables as reported in their respective publicly available datasets (e.g. no conversions).

### 2.3. Statistical analyses

To evaluate the diagnostic potential of easy-to-obtain clinical measures for dementia, we used several machine learning algorithms including tree-based algorithms (classification and regression trees (CART), random forest, gradient boosting machines (GBM), and extreme gradient boosting (XGBoost) and artificial neural networks. Datasets (ADNI only, AIBL only, or a combined ADNI and AIBL) were split into a 70% training dataset and a 30% held-out testing dataset. Machine learning models were built, fine-tuned, and validated on the training datasets using five-fold cross-validation repeated five times. Where required due to class imbalances of the output variable, an oversampling technique was used. Here, the underrepresented class was randomly resampled to ensure that the algorithms received approximately the same number of classes. For all algorithms, a fine-tuning grid method was used where we estimated all possible combinations of parameters within the predetermined ranges (see Supplementary Table 1 for hyperparameters). All analyses were done in R (version 4.3.1, R Core Team, 2023) using ‘caret’ package [22].

### 2.4. Model evaluation

The performance of the machine learning models was evaluated using a 30% held-out dataset. For each model, we report several metrics including: sensitivity (correctly identified dementia cases), positive predictive value (PPV also known as precision; number of dementia cases / total number of predicted dementia cases (true and false)), specificity (correctly identified healthy controls), negative predictive value (NPV; number of healthy controls / total number of predicted healthy controls (true and false)), and AUC (ability to distinguish dementia cases and healthy controls). The main performance indicators we used in the present study were sensitivity and PPV. Using these metrics is important because it reduces the likelihood of a dementia case being identified as a healthy control (false negative) and provides the probability that a person with a positive result indeed has dementia [23]. Further, as shown by others, multifactorial dementia prediction models with a high level of specificity in the absence of sensitivity are unable to identify the majority of dementia cases [18]. Where sensitivities and PPVs were similar (<0.2 difference) between two models, we also considered specificity and NPV when identifying the top performing model, as we acknowledge that dementia prediction models still need reasonable metrics for identifying true healthy controls.

To evaluate the relative contribution of features to overall performance of our models we performed a SHAP (SHapley Additive exPlanations) analysis. This feature importance selection method allowed us to assign an importance value for each input variable (feature) for our predictions [24], thus demonstrating which variables are the most important for dementia prediction. SHAP was done in R using package ‘shapviz’.

## 3. RESULTS

### 3.1. APOE genotype shows the highest utility for a dementia diagnosis using a merged ADNI-AIBL cohort

We first merged the ADNI and AIBL cohorts into a single large dataset to test the diagnostic potential of 21 easy-to-obtain clinical measures for dementia. This merged dataset was then randomly split into a 70% training and validation set and a 30% unseen testing dataset. With APOE genotype included as a feature, model sensitivity ranged from 0.63 to 0.83 and PPV ranged from 0.69 to 0.82 (Table 2). The neural network had the highest sensitivity of 0.83 and a PPV of 0.73 with a specificity of 0.66 and NPV of 0.78. Specificity and NPV was generally higher than sensitivity and PVV, ranging from 0.66 to 0.86 and 0.73 to 0.78, respectively (Table 2). To identify which of the 21 features (clinical measures) were contributing the most to our models’ performance, we used a SHAP analysis. This showed that APOE genotype had the highest contribution to our models’ ability to predict a dementia diagnosis (Figure 1A,B). Further, the high positive SHAP value indicated that APOE genotype was the most valuable for identifying a dementia patient (sensitivity and PPV; Figure 1B). The urea nitrogen test (RCT6), was shown to be a second, albeit lesser, contributor to model performance (Figure 1A,B). Unlike APOE genotype, urea nitrogen was more important for identifying a healthy control (specificity and NPV; Figure 1B). Importantly, the urea nitrogen finding may be consequence of differences in the median between ADNI and AIBL (approximately 18 and 35, respectively). We were unable to determine if this may be reflective of unit of measure differences between ADNI and AIBL, as these data were not reported.

**Figure 1.**
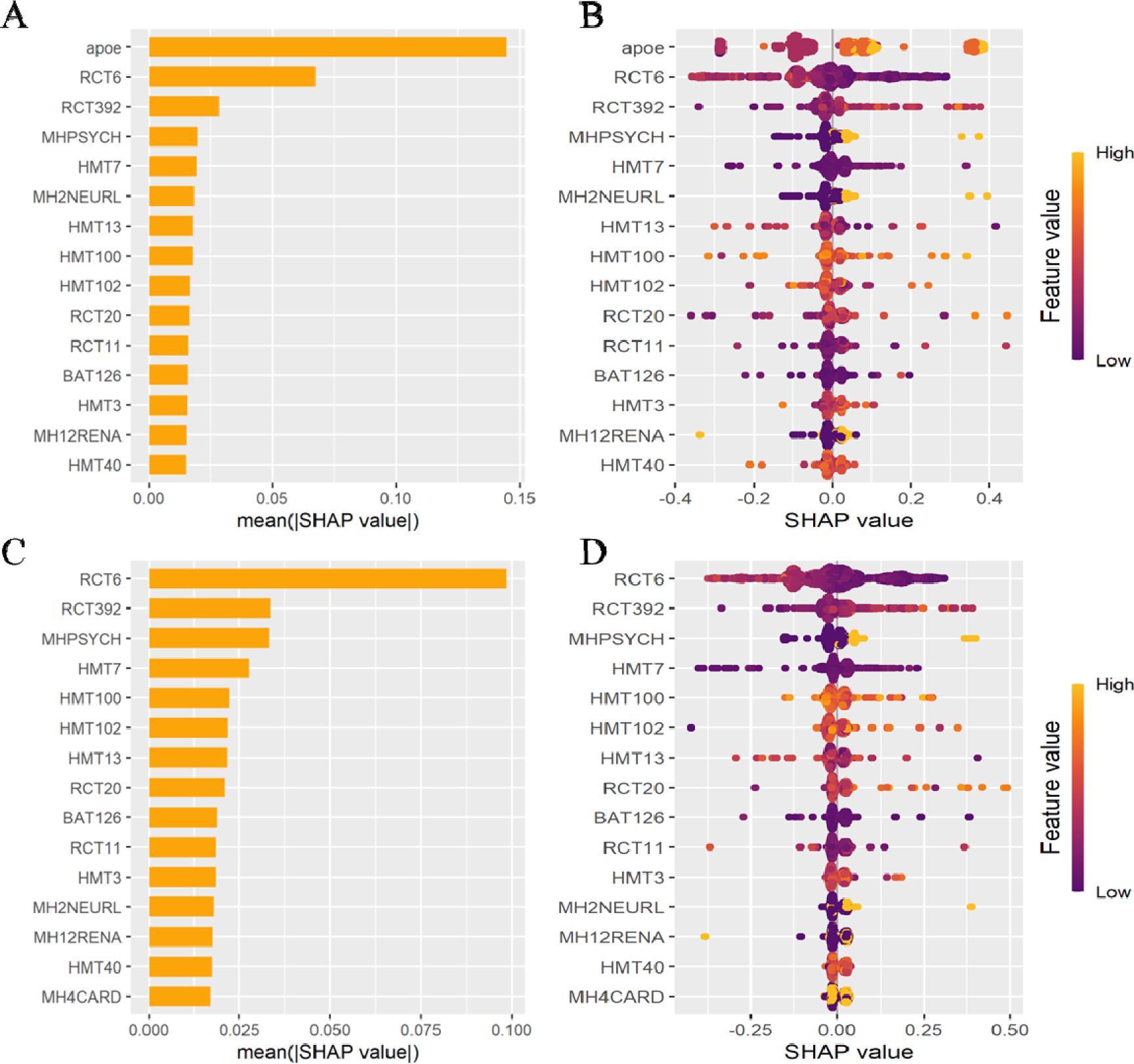
Feature importance using SHAP analysis in our models. (A,C) Absolute SHAP value for the top 15 features in models with and without APOE genotype, respectively. (B,D) Heat map of relative contribution of each feature in models with and without APOE genotype, respectively. Negative values = healthy control and positive values = dementia. Abbreviations: BAT126: vitamin B12; HMT3: red blood cell count; HMT7: white blood cell count; HMT13: platelets; HMT40: hemoglobin; HMT100: mean corpuscular hemoglobin; HMT102: mean corpuscular hemoglobin concentration; MH2NEURL: medical history neurologic; MH4CARD: medical history cardiovascular; MH12RENA: medical history renal-genitourinary; MHPSYCH: medical history psychiatric; RCT6: urea nitrogen; RCT11: serum glucose; RCT20: cholesterol (high performance); RCT392: creatinine.

**Table 2.** Diagnostic performance of algorithms on a unified ADNI-AIBL dataset with and without APOE genotype as a feature.

|  | **Sensitivity** |  | **Specificity** |  | **PPV (precision)** |  | **NPV** |  | **AUC** |  |
| --- | --- | --- | --- | --- | --- | --- | --- | --- | --- | --- |
|  | APOE | No APOE | APOE | No APOE | APOE | No APOE | APOE | No APOE | APOE | No APOE |
| Classification and regression trees (CART) | 0.71 | 0.57 | 0.85 | 0.79 | 0.81 | 0.76 | 0.77 | 0.61 | 0.84 | 0.68 |
| Random forest | 0.69 | 0.58 | 0.86 | 0.77 | 0.82 | 0.71 | 0.75 | 0.66 | 0.87 | 0.75 |
| Shrinkage discriminant analysis (SDA) | 0.68 | 0.61 | 0.85 | 0.80 | 0.82 | 0.75 | 0.74 | 0.67 | 0.84 | 0.76 |
| General linear model | 0.67 | 0.61 | 0.84 | 0.81 | 0.79 | 0.76 | 0.74 | 0.67 | 0.84 | 0.77 |
| K nearest neighbour | 0.63 | 0.61 | 0.78 | 0.75 | 0.69 | 0.64 | 0.73 | 0.73 | 0.68 | 0.72 |
| Gradient boosting machines (GBM) | 0.72 | 0.61 | 0.85 | 0.81 | 0.81 | 0.76 | 0.74 | 0.67 | 0.87 | 0.75 |
| Extreme gradient boosting (XGBoost) | 0.70 | 0.59 | 0.85 | 0.81 | 0.81 | 0.77 | 0.77 | 0.65 | 0.86 | 0.75 |
| Neural network | 0.83 | 0.73 | 0.66 | 0.60 | 0.73 | 0.65 | 0.78 | 0.79 | 0.75 | 0.72 |

To further demonstrate that APOE genotype is essential for the models’ predictive performance, we repeated all analyses but excluded APOE genotype from our selected features (20 features instead of 21). As expected, performance significantly decreased across all models (Table 2). Without APOE genotype, sensitivity and PPV ranged from 0.57 to 0.73 and 0.64 to 0.77, respectively. Although specificity remained relatively high, ranging from 0.60 to 0.81, NPV generally decreased across the models, with a range of 0.61 to 0.79. Urea nitrogen took the place of APOE genotype as the top contributor to model performance (Figure 1C,D). As before, urea nitrogen was more important for identifying a healthy control (specificity and NPV; Figure 1D), suggesting that this may contribute to the higher specificity of our models without APOE genotype. Though again, it’s unclear if this is due to median differences between ADNI and AIBL.

### 3.2. Multivariate dementia prediction models perform poorly when using either the ADNI or AIBL datasets as training and testing

To further identify if our multivariate prediction models of dementia are generalizable, we separated the dataset back into the ADNI and AIBL cohorts and re-tested our models. In the first experiment, we trained and validated our 21-feature models (including APOE genotype) on ADNI and then tested them on AIBL and, in the second, we performed the reverse. When we trained our models using the ADNI dataset, they lost dementia diagnostic utility. Here, our sensitivity substantially decreased to a range of 0.25 to 0.36 however our PPV remained higher and ranged from 0.62 to 0.78 (Table 3). Specificity, on the other hand, was very high with a range of 0.90 to 0.93, similar to existing dementia diagnostic models. NPV was lower and highly variable, ranging from 0.47 to 0.97 (Table 3). There were two top performing models: CART and GBM. Both had sensitivities of 0.35, PPVs of 0.72, specificities of 0.93 and NPVs of 0.73.

**Table 3.** Diagnostic performance of algorithms using opposite training and testing datasets ((**A**) ADNI train, AIBL test; or (**B**) AIBL train, ADNI test)

|  | Sensitivity |  | Specificity |  | PPV (precision) |  | NPV |  | AUC |  |
| --- | --- | --- | --- | --- | --- | --- | --- | --- | --- | --- |
|  | A | B | A | B | A | B | A | B | A | B |
| Classification and regression trees (CART) | 0.35 | 0.83 | 0.93 | 0.61 | 0.72 | 0.71 | 0.73 | 0.76 | 0.73 | 0.73 |
| Random forest | 0.27 | 0.81 | 0.92 | 0.39 | 0.74 | 0.80 | 0.59 | 0.97 | 0.72 | 0.70 |
| Shrinkage discriminant analysis (SDA) | 0.29 | 0.84 | 0.93 | 0.49 | 0.77 | 0.45 | 0.61 | 0.86 | 0.75 | 0.70 |
| General linear model | 0.29 | 0.61 | 0.93 | 0.37 | 0.78 | 0.51 | 0.62 | 0.47 | 0.75 | 0.58 |
| K nearest neighbour | 0.25 | 0.70 | 0.90 | 0.47 | 0.68 | 0.60 | 0.58 | 0.59 | 0.68 | 0.63 |
| Gradient boosting machines (GBM) | 0.35 | 0.83 | 0.93 | 0.61 | 0.72 | 0.71 | 0.73 | 0.76 | 0.73 | 0.73 |
| Extreme gradient boosting (XGBoost) | 0.36 | 0.80 | 0.93 | 0.58 | 0.72 | 0.68 | 0.73 | 0.73 | 0.74 | 0.74 |
| Neural network | 0.29 | 0.89 | 0.93 | 0.39 | 0.62 | 0.99 | 0.77 | 0.50 | 0.70 | 0.51 |

Interestingly, training our models on the AIBL dataset and testing them on the ADNI dataset improved their predictive power. In this instance, sensitivity and PPV improved to a range of 0.61 to 0.89 and 0.45 to 0.99, respectively (Table 3). Our specificity and NPV, however, decreased to a range of 0.37 to 0.61 and 0.47 to 0.97, respectively (Table 3). CART and GBM were again the equally the top performing models with sensitivities of 0.83, specificities of 0.61, PPVs of 0.71, and NPVs of 0.76. Combined, this suggests that almost all the models, except CART and GBM, have a poor predictive performance when separating the ADNI and AIBL cohorts.

### 3.3. The relative distributions of APOE genotype across ADNI and AIBL drive poor model performance

We next sought out to better understand why our CART and GBM models had an acceptable level of performance when training and testing on the opposite datasets. To do this, we chose to look at CART performance, specifically, as we can use decision trees to better understand the features used for class identification. The CART decision tree for both experiments (train ADNI, test AIBL and train AIBL, test ADNI) was identical and showed APOE genotype was the only feature that it was using to predict whether a participant was a healthy control or a dementia patient (Figure 2). If the APOE genotype contained at least one APOE4 allele (e4,e2 or e4,e3 or e4,e4) then the models categorized that person as having dementia and, if not (i.e. an APOE genotype of e2,e2 or e2,e3 or e3,e3), then as a healthy control.

**Figure 2.**
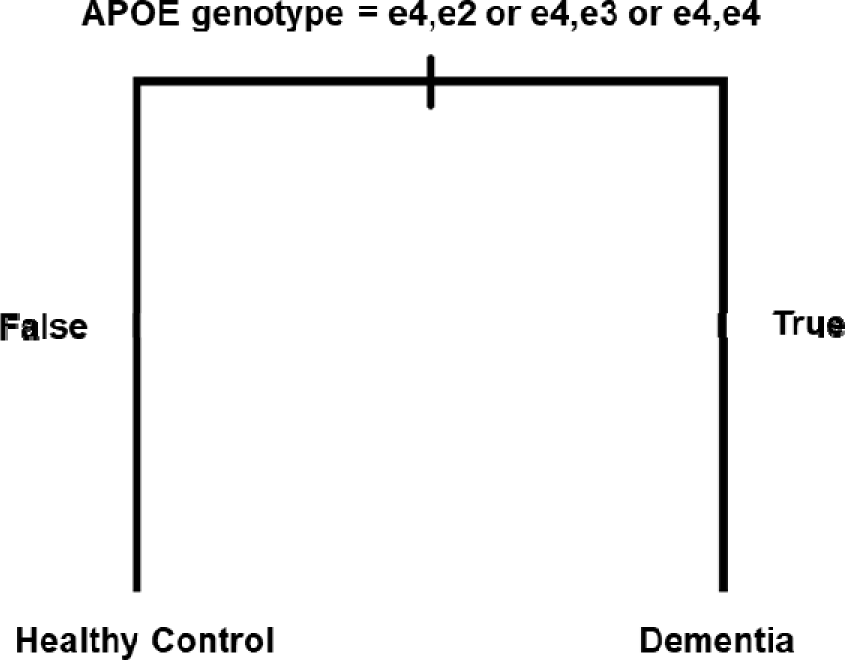
Decision tree used by the classification and regression tree (CART) models when trained on ADNI and tested on AIBL or trained on AIBL and tested on ADNI.

Despite both of our CART models using only APOE genotype to determine a healthy control from a dementia patient, it was unclear why performance differed dramatically depending on if we used ADNI or AIBL as the training or testing dataset (Table 2). Specifically, our metrics in Table 2 showed that the AIBL tested model had a high rate of false positives and the ADNI tested model had a higher rate of false negatives. This suggested that a difference in the relative distribution of APOE genotypes in healthy controls and dementia patients across the ADNI and AIBL cohorts may be driving performance differences. To confirm that this was the case, we examined the relative distribution of APOE genotypes in healthy controls and dementia patients in ADNI and AIBL. As shown in Figure 3A, AIBL indeed has a high rate of false positives – i.e. the CART misclassified APOE4 allele carriers as having dementia when they were instead healthy controls. ADNI, on the other hand, had a slightly higher rate of false negatives than AIBL (Figure 3B), but only for the e3,e2 genotype. Here, the CART misclassified these as being healthy controls when they had dementia.

**Figure 3.**
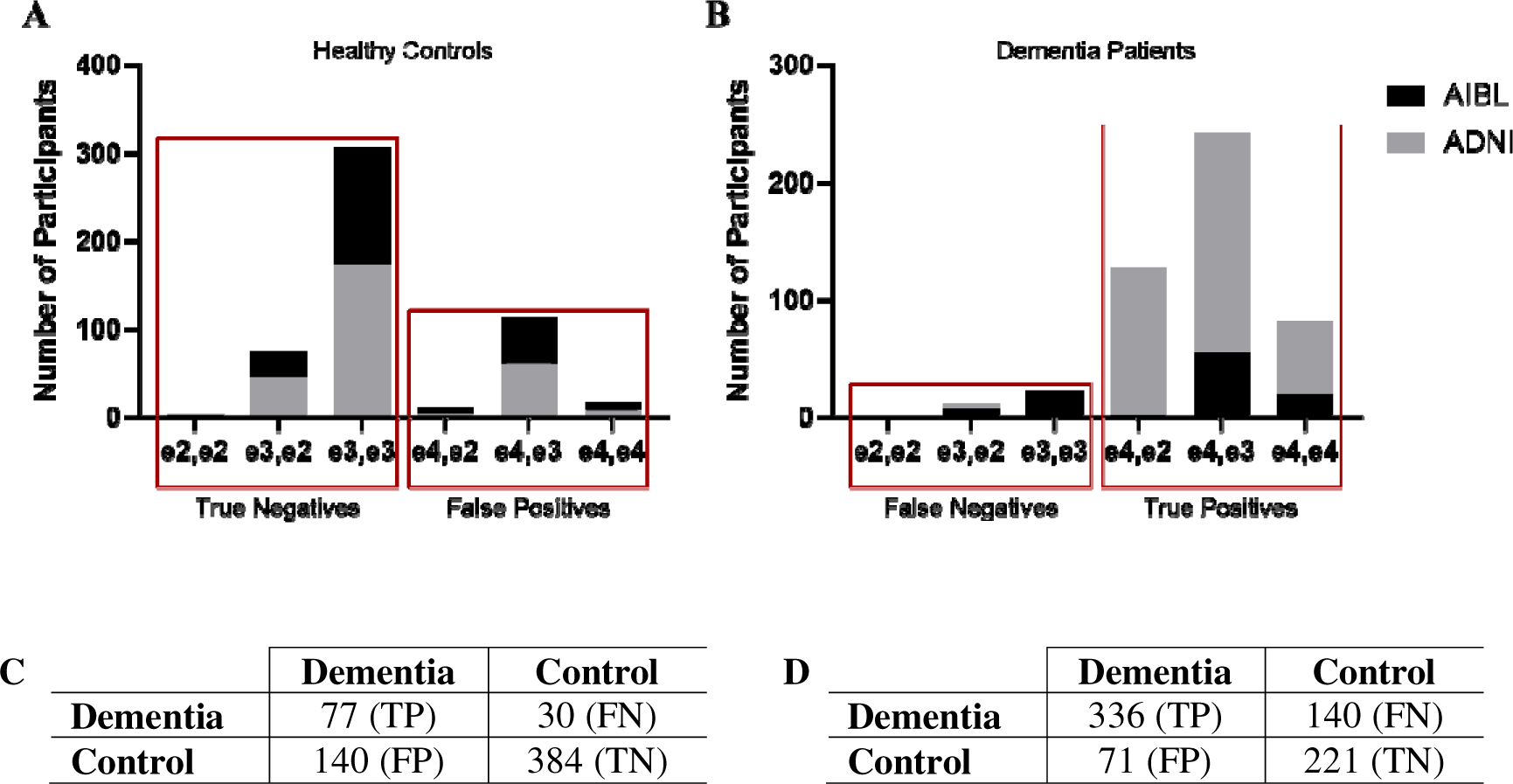
Misclassifications in classification and regression tree (CART) models. (A-B) Relative distribution of APOE genotypes across the ADNI and AIBL datasets for (A) healthy controls and (B) dementia patients. Red boxes highlight the number of true negatives and positives and false negatives and positives across the groups. (C-D) Confusion matrices for CART models when (C) trained on ADNI and tested on AIBL and (D) trained on AIBL and tested on ADNI. Abbreviations: FN: false negative; FP: false positive; TN: true negative; FN: false negative.

We further confirmed this using the CART confusion matrices. Indeed, when our CART models were tested on AIBL there was a high rate of false positives (i.e. low sensitivity; Figure 3C). When they were tested on ADNI, however, there was a higher rate of false negatives (i.e. lower specificity; Figure 3D).

### 3.4. APOE genotype predicts dementia cases in ADNI whereas it predicts healthy controls in AIBL

To further investigate the basis for our models’ poor predictive performance when training and testing on opposite datasets, we examined the predictive capabilities within ADNI and AIBL independently. First, we examined the performance of our 21-feature (including APOE genotype) models when we trained and validated on a 70% split of the ADNI dataset and tested on a withheld 30% of the ADNI dataset. Our models showed strong dementia predictive performance. Here, depending on the algorithm, our models showed a very high sensitivity ranging from 0.80 to 0.86 and a PPV ranging from 0.65 to 0.75 (Table 4). The two models with the highest performance were the CART and XGBoost with an equal sensitivity of 0.86 and PPV of 0.84 (Table 4). Our SHAP analysis again demonstrated that APOE genetic testing was the main feature driving our predictive power for identifying incident dementia cases (Figure 4A,B).

**Figure 4.**
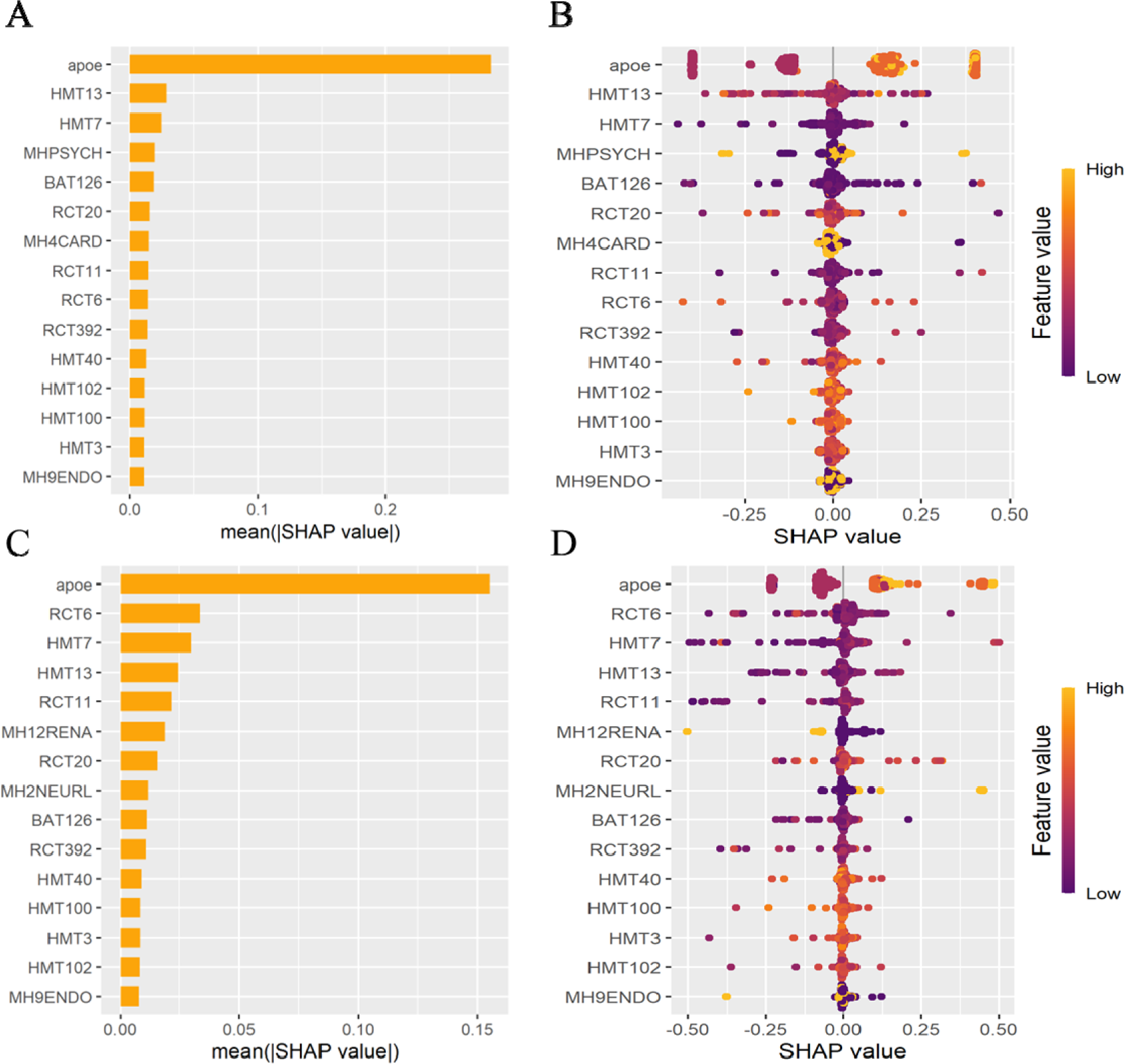
Feature importance using SHAP analysis in independent ADNI or AIBL models. (A-B) SHAP analysis for ADNI models showing (A) absolute SHAP value for the top 15 features and (B) heat map of the relative contribution of each feature. (C-D) SHAP analysis for AIBL models showing (C) absolute SHAP value for the top 15 features and (D) heat map of the relative contribution of each feature. Negative values = healthy control, positive values = dementia. Abbreviations: BAT126: vitamin B12; HMT3: red blood cell count; HMT7: white blood cell count; HMT13: platelets; HMT40: hemoglobin; HMT100: mean corpuscular hemoglobin; HMT102: mean corpuscular hemoglobin concentration; MH2NEURL: medical history neurologic; MH4CARD: medical history cardiovascular; MH12RENA: medical history renal-genitourinary; MHPSYCH: medical history psychiatric; RCT6: urea nitrogen; RCT11: serum glucose; RCT20: cholesterol (high performance); RCT392: creatinine.

**Table 4.** Diagnostic performance of machine learning algorithms for either the ADNI or AIBL datasets, respectively.

|  | **Sensitivity** |  | **Specificity** |  | **PPV (precision)** |  | **NPV** |  | **AUC** |  |
| --- | --- | --- | --- | --- | --- | --- | --- | --- | --- | --- |
|  | **ADNI** | **AIBL** | **ADNI** | **AIBL** | **ADNI** | **AIBL** | **ADNI** | **AIBL** | **ADNI** | **AIBL** |
| Classification and regression trees (CART) | 0.86 | 0.35 | 0.67 | 0.94 | 0.74 | 0.85 | 0.82 | 0.59 | 0.78 | 0.68 |
| Random forest | 0.84 | 0.40 | 0.65 | 0.94 | 0.71 | 0.81 | 0.79 | 0.68 | 0.80 | 0.79 |
| Shrinkage discriminant analysis (SDA) | 0.86 | 0.41 | 0.67 | 0.90 | 0.72 | 0.67 | 0.82 | 0.75 | 0.77 | 0.79 |
| General linear model | 0.84 | 0.42 | 0.64 | 0.88 | 0.71 | 0.59 | 0.80 | 0.79 | 0.78 | 0.77 |
| K nearest neighbour | 0.80 | 0.26 | 0.59 | 0.87 | 0.65 | 0.74 | 0.75 | 0.45 | 0.72 | 0.68 |
| Gradient boosting machines (GBM) | 0.86 | 0.42 | 0.68 | 0.95 | 0.74 | 0.85 | 0.82 | 0.69 | 0.80 | 0.79 |
| Extreme gradient boosting (XGBoost) | 0.86 | 0.43 | 0.68 | 0.92 | 0.74 | 0.84 | 0.82 | 0.68 | 0.82 | 0.79 |
| Neural network | 0.84 | 0.44 | 0.68 | 0.86 | 0.75 | 0.56 | 0.79 | 0.72 | 0.77 | 0.68 |

We then performed the same experiment, however this time using 70% of the AIBL dataset to train and validate our models and a withheld 30% of AIBL to test them. These models were unable to predict dementia cases. Here, the sensitivity ranged from 0.26 to 0.44 (Table 4), suggesting that the models miss most dementia cases. The PPV, however, was higher, ranging from 0.56 to 0.85 (Table 4). Despite this, and unlike the ADNI dataset, our models had very high specificity that ranged from 0.86 to 0.95 (Table 4). The NPV ranged from 0.45 to 0.75 (Table 4). Again, APOE genotype remained the strongest predictor that drove model performance (Figure 4C,D) and urea nitrogen testing came in as a second predictor, albeit to a lesser extent than APOE genotype.

Combined, these findings indicate that APOE genotype is predictive of dementia cases in the ADNI cohort (high sensitivity) whereas it’s predictive of healthy controls in the AIBL cohort (high specificity).

## 4. DISCUSSION

Developing diagnostic models of dementia that rely on “easy-to-obtain” clinical measures rather than neuroimages is important for timely diagnoses by primary care physicians. Using cohort data from the ADNI and AIBL datasets, this study used machine learning to examine the diagnostic potential of 21 clinical measures, including APOE genotype, medical history, hematological and other blood tests.

Using a combined ADNI and AIBL cohort dataset, we showed that artificial neural networks have the best predictive performance. Our sensitivity was 0.83 and PPV was 0.73, suggesting that our neural network would only miss approximately 27% of incident dementia cases. These metrics showed that our neural network outperformed existing models [9, 14, 16-18] and that it may hold practical utility for dementia diagnostics. APOE genotype was the top performing variable with urea nitrogen the second, albeit lesser, performing variable.

Interestingly, when we separated the ADNI and AIBL cohorts and used them either as training or testing datasets, respectively, our models lost predictive power. When we trained our models on ADNI and tested them on AIBL, our models’ sensitivity decreased to a clinically invaluable range of 0.25-0.36. When we performed the reverse, training on AIBL and testing on ADNI, our sensitivity metrics improved however our specificity fell to 0.37-0.61. We identified that these discrepancies were due to differences in the relative distribution of APOE genotype across ADNI and AIBL. ADNI had a small increase in the number of false negatives due to APOE4 non-carriers being identified as healthy controls when they were instead dementia cases. AIBL, on the other hand, had a very high number of false positives because APOE4 carriers were being classified as having dementia when they were healthy controls. Therefore, although APOE genotype was the main predictive variable across all our models, APOE genotype better predicted dementia cases in ADNI whereas it was predictive of healthy controls in AIBL.

The reasons for this discrepancy in APOE genotype are largely unclear. It may be due to class imbalances across the datasets: ADNI had more dementia cases whereas AIBL had more healthy controls. This may have led to higher sensitivity using ADNI and a higher specificity when using AIBL. It may also be due to differences in participant group allocations between ADNI and AIBL. ADNI reported that participants with a subjective memory complaint were placed into the dementia group whereas AIBL reported that these participants went into the healthy control group [19, 20]. As reported in the baseline and methodology characteristics study, this resulted in approximately 50% of the healthy control group in AIBL made up of participants that had a subjective memory complaint – i.e. said yes in response to being asked “Do you have difficulties with your memory” [20]. Subjective memory complaints are an important consideration in the context of APOE genotype. Numerous studies have identified that APOE4 carriers with subjective memory complaints or cognitive dysfunction have worse baseline memory [25], memory-guided attention [26], and episodic memory [27]. Age has also been shown to play a significant role in the relationship between APOE4 and a faster rate of memory decline [25, 26]. Further, some studies have shown that APOE4 carriers with subjective memory complaints and cognitive dysfunction are similar to people with early mild cognitive impairment (MCI) [28] and are more likely to develop clinical MCI [29, 30]. People with subjective memory complaints and an APOE4 allele also have changes in the brain that are indicative of MCI and dementia including hippocampal volume changes [27, 31]. One study even found evidence to suggest that subjective memory complaints may be a realistic appraisal of cognitive decline-associated brain changes in people with and APOE4 allele [31]. It may be the case, therefore, that AIBL contains healthy controls, especially those with an APOE4 allele, that are in the early stages of MCI or dementia.

It’s important to note that the finding that our AIBL-derived models have a high rate of false positives may be due to the data that has been made open source and publicly available. For example, a study using a subgroup of participants from the AIBL cohort found that subjective memory complaints were indicative of a higher amyloid β burden and APOE4 carrier status [32]. The subjective memory complaint status of the participants in the publicly available AIBL cohort is not available. We were unable, therefore, to further investigate if re-coding the subjective memory complainers as dementia cases led to improved model performance in line with what we found in our ADNI-based models. Future research would benefit from further examining this.

Large open source and publicly available dementia datasets are increasingly becoming available and, in line with this, it is becoming easier to pool participants to increase study sample sizes and improve statistical power. This is particularly important for studies like ours that use machine learning or other artificial intelligence-based methods to unravel complex relationships between variables for dementia diagnostic and treatment modelling. Our results, however, highlight several limitations of using such cohort data. First, there needs to be increased effort in ensuring that the data obtained between cohorts is the same. For example, although the test results of the hematological and other blood tests largely looked to be similar across ADNI and AIBL, there were clear differences in the urea nitrogen results. This is likely due to differences in the units of measurement used (e.g. ng/dL vs. nmol/L) however without this information it’s not possible to confirm. Unclear units of measurement also may limit the clinical implementation of these types of models and their predictive features. There were also many variables missing between the cohorts that may be important. For example, ADNI included measures like ethnicity, educational attainment, and type of residence that were not including in AIBL, thus potentially limiting the generalizability of our models (e.g. to multiple ethnicities). Second, our results highlight that it’s important to independently examine the behaviour of different dementia cohorts prior to undertaking studies using merged or unified datasets. Here, our merged ADNI-AIBL models performed very well, despite there being clear and opposing differences when the cohorts were examined individually. It’s not clear, therefore, whether the merging of the two datasets produced a cohort that is truly generalizable to the population. For example, merging datasets may lead to the unintentional washing out of important variables and effects that are relevant to dementia. This may limit the interpretability and generalizability of studies that only use merged cohorts and hold important implications for future research on predictive dementia models.

Irrespective of the differences between the ADNI and AIBL cohorts, APOE genotype remained the strongest predictor of both dementia cases and healthy controls. In line with the growing body of research, this finding highlights the importance of considering APOE genotype in the context of dementia diagnostics. APOE genetic testing is a low-cost and readily available assay. In the US, an APOE genetic screen costs from $100-$125 USD and can even be done from home and mailed into a diagnostic testing lab. Similarly, APOE testing in Australia costs $150-$200 AUD and can be done through routine pathology labs. Although the UK’s National Health Service (NHS) doesn’t offer APOE testing, there are services available offering APOE genetic screens in the context of cardiovascular disease for similar price ranges (£180). Further, APOE genetic results are easy to interpret in the absence of specialist resources. This highlights that it’s a practical diagnostic tool that primary care physicians can readily use to identify patients with subjective memory complaints who are at a high risk of dementia. Future research, therefore, should examine the use of routine APOE genetic testing in primary care clinics for the purposes of early identification and diagnosis of dementia.

## 5. CONCLUSION

In conclusion, we have identified that across several easy-to-obtain clinical measures, APOE genotype remains the best predictor of dementia cases relative to healthy controls. APOE genotype remains a relatively widely available test and consideration should be given to its utility as a routine diagnostic test for dementia. We also highlighted that there are limitations associated with using publicly available cohort data to generate multivariate dementia prediction models. These limitations warrant further efforts to unify existing and future dementia cohorts.

## Data Availability

All data produced in the present study are available upon reasonable request to the authors

## ACKNOWLEDGMENTS

The authors are grateful to the Alzheimer’s Disease Neuroimaging Initiative (ADNI) and the Australian Imaging, Biomarkers, and Lifestyle Study of Ageing (AIBL) studies for providing the data. This research did not receive any specific grant from funding agencies in the public, commercial, or not-for-profit sectors. C.A.F. is supported by philanthropic funding from the Leece Family Foundation and the Neil & Norma Hill Foundation. Open access funding provided to CAF by the Westmead Institute for Medical Research.

## CONSENT STATEMENT

All participants of the Alzheimer’s Disease Neuroimaging Initiative (ADNI) and the Australian Imaging, Biomarkers, and Lifestyle Study of Ageing (AIBL) studies provided informed consent.

## REFERENCES

[1] World Health Organization. Dementia. 2023.

[2] Liss JL, Assuncao SS, Cummings J, Atri A, Geldmacher DS, Candela SF, et al. Practical recommendations for timely, accurate diagnosis of symptomatic Alzheimer’s disease (MCI and dementia) in primary care: A review and synthesis. Journal of Internal Medicine. 2021;290:310–34.

[3] Parker M, Barlow S, Hoe J, Aitken L. Persistent barriers and facilitators to seeking help for a dementia diagnosis: A systemtic review of 30 years of the perspectives of carers and people with dementia. International Psychogeriatrics. 2020;32:611-34.

[4] Pellegrini E, Ballerini L, Hernandez MCV, Chappell FM, Gonzalez-Castro V, Anblagan D, et al. Machine learning of neuroimaging for assisted diagnosis of cognitive impairment and dementia: A systematic review. Alzheimer’s & Dementia: Diagnosis, Assessment & Disease Monitoring. 2022;10:519–35.

[5] Javeed A, Dallora AL, Berglund JS, Ali A, Ali L, Anderberg P. Machine learning for dementia prediction: A systematic review and future research directions. Journal of Medical Systems. 2023;47.

[6] Leming MJ, Bron EE, Bruffaerts R, Ou Y, Iglesias JE, Gollub RL, et al. Challenges of implementing computer-aided diagnostic models for neuroimages in a clinical setting. npj Digital Medicine. 2023;6.

[7] Mansfield E, Noble N, Sanson-Fisher R, Mazza D, Bryant J. Primary care physicians’ perceived barriers to optimal demenetia care: A systematic review. The Gerontologist. 2018;59:697-708.

[8] Alzheimer’s Association. Alzheimer’s Association facts and figures. Alzheimer’s & Dementia. 2020:391–460.

[9] Kivipelto M, Ngandu T, Laatikainen T, Winblad B, Soininen H, Tuomilehto J. Risk score for the prediction of dementia risk in 20 years among middle aged people: A longitudinal, population-based study. The Lancet Neurology. 2006;5:735–41.

[10] Luck T, Riedel-Heller SG, Luppa M, Wiese B, Wollny A, Wagner M, et al. Risk factors for incident mild cognitive impairment - Results from the German study on ageing, cognition and dementia in primary care patients (AgeCoDe). Acta Psychiatrica Scandinavica. 2010;121:260–72.

[11] Anstey KJ, Cherbuin N, Herath PM, Qiu C, Kuller LH, Lopez OL, et al. A self-report risk index to predict occurrence of dementia in three independent cohorts of older adults: The ANU-ADRI. PLoS One. 2014;9:e86141.

[12] Capuano AW, Shah RC, Blanche P, Wilson RS, Barnes LL, Bennett D, et al. Derivation and validation of the rapid assessment of dementai risk (RADaR) for older adults. PLoS One. 2022;17:e0265379.

[13] Barnes DE, Beiser AS, Lee A, Langa KM, Koyama A, Preis SR, et al. Development and validation of a brief dementia screening indicator for primary care. Alzheimer’s & Dementia. 2014;10:656–65.

[14] James C, Ranson JM, Everson R, Llewellyn DJ. Performance of machine learning algorithms for predicting progression to dementia in memory clinic patients. JAMA Network Open. 2021;4:e2136553.

[15] So A, Hooshyar D, Park KW, Lim HS. Early diagnosis of dementia from clinical data by machine learning techniques. Applied Sciences. 2017;7:651.

[16] Jessen F, Wiese B, Bickel H, Eifflander-Gorfer S, Fuchs A, Kaduszkiewicz H, et al. Prediction of dementia in primary care patients. PLoS One. 2011;6:e16852.

[17] Walters K, Hardoon S, Petersen I, Iliffe S, Omar RZ, Nazareth I, et al. Dementia risk score using routinely collected data. BMC Medicine. 2016;14.

[18] Kivimaki M, Livingston G, Singh-Manoux A, Mars N, Lindbohm JV, Pentti J, et al. Estimating dementia risk using multifactorial prediction models. JAMA Network Open. 2023;6:e2318132.

[19] Petersen RC, Aisen PS, Beckett LA, Donohue MC, Gamst AC, Harvey DJ, et al. Alzheimer’s Disease Neuroimaging Initiative (ADNI): Clinical characterization. Neurology. 2009;74:201–9.

[20] Ellis KA, Bush AI, Darby D, De Fazio D, Foster J, Hudson P, et al. The Australian Imaging, Biomarkers and Lifestyle (AIBL) study of aging: Methodology and baseline characteristics of 1112 individuals recruited for a longitudinal study of Alzheimer’s disease. International Psychogeriatrics. 2009;21:672–87.

[21] McKahnn G, Drachman D, Folstein M, Katzman R, Price D, Stadlan EM. Clinical diagnosis of Alzheimer’s disease: Report of the NINCDS-ADRDA work group under the auspices of Department of Health and Human Services Task Force on Alzheimer’s Disease. Neurology. 1984;34:939–44.

[22] Kuhn M. Building predictive models in R using the caret package. Journal of Statistical Software. 2008;28:1–26.

[23] Trevethan R. Sensitivity, specificity, and predictive values: Foundations, pliabilities, and pitfalls in research and practice. Frontiers in Public Health. 2017;5.

[24] Lundberg SM, Lee SI. A unified appraoch to interpreting model predictions. 31st Conference on Neural Inforamtion Processing Systems (NIPS). Long Beach, California 2017.

[25] Samieri C, Proust-Lima C, Glymour MM, Okereke OI, Amariglio RE, Sperling RA, et al. Subjective cognitive concerns, episodic memory, and the APOE ε4 allele. Alzheimer’s & Dementia. 2014;10:752–9.

[26] Zimmerman J, Alain C, Butler C. Impaired memory-guided attention in asymptomatic APOE4 carriers. Scientific Reports. 2019;9.

[27] Striepens N, Scheef L, Wind A, Meiberth D, Popp J, Spottke A, et al. Interaction effects of subjective memory impairment and APOE4 genotype on episodic memory and hippocampal volume. Psychological Medicine. 2011;41.

[28] Cho H, Kim Y-E, Chae W, Kim KW, Kim J-W, Kim HJ, et al. Distribution and clinical impact of apolipoprotein E4 in subjective memory impairment and early mild cognitive impairment. Scientific Reports. 2020;10.

[29] Muller-Gerards D, Weimar C, Abramowski J, Tebrugge S, Jokisch M, Dragano N, et al. Subjective cognitive decline, APOE ε4, and incident mild cognitive impairment in men and women. Alzheimer’s & Dementia. 2019;11:221–30.

[30] Ali JI, Smart CM, Gawryluk JR. Subjective cognitive decline and APOE ε4: A systematic review. Journal of Alzheimer’s Disease. 2018;65:303–20.

[31] Stewart R, Godin O, Crivello F, Maillard P, Mazoyer B, Tzourio C, et al. Longitudinal neuroimaging correlates of subjective memory impairment: 4-year prospective community study. The British Journal of Psychiatry. 2011;198:199–205.

[32] Zwan MD, Villemagne V, Dore V, Buckley R, Bourgeat P, Veljanoski R, et al. Subjective memory complaints in APOE ε4 carriers are associated with high amyloid-β burden. Journal of Alzheimer’s Disease. 2015;49:1115–22.

