## Supplementary Table for "Developing multifactorial dementia prediction models using clinical variables from cohorts in the US and Australia"

7. Data used in preparation of this article were obtained from the Alzheimer’s Disease Neuroimaging Initiative (ADNI) database (adni.loni.usc.edu). As such, the investigators within the ADNI contributed to the design and implementation of ADNI and/or provided data but did not participate in analysis or writing of this report. A complete listing of ADNI investigators can be found at: <http://adni.loni.usc.edu/wp-content/uploads/how_to_apply/ADNI_Acknowledgement_List.pdf>

8. Data used in the preparation of this article was obtained from the Australian Imaging Biomarkers and Lifestyle flagship study of ageing (AIBL) funded by the Commonwealth Scientific and Industrial Research Organisation (CSIRO) which was made available at the ADNI database (www.loni.usc.edu/ADNI). The AIBL researchers contributed data but did not participate in analysis or writing of this report. AIBL researchers are listed at [www.aibl.csiro.au](http://www.aibl.csiro.au).

\# These authors contributed equally to the work.

**Supplementary Table 1.** Hyperparameters used for machine learning models

| **Model** | **Parameters** |
| --- | --- |
| Linear regression | α: 0, 1  λ: 0.001-1 |
| Shrinkage discriminant analysis (SDA) | Diagonal: true, false  λ: 0.001-1 |
| K-nearest neighbour (KNN) | Number of the nearest neighbours: 1-15 |
| Classification and regression trees (CART) | Complexity parameter: 0.0001-0.2 |
| Random forest | Number of randomly selected predictors: 1-# of features in dataset |
| GBM | Number of boosting iterations: 1-1000,  Maximum tree depth: 1:100,  η: 0.00001:0.1  Minimum terminal node size: 1:50 |
| XGBoost | Number of boosting iterations: 1-1000,  Maximum tree depth: 1:100,  η: 0.00001:0.1,  Minimum loss reduction: 0.05-3,  Subsample ratio of columns: 0.1-5  Minimum size of instance weight: 1:30  Subsample percentage: 1:50 |
| Neural network | Number of hidden layers: 0 and 1  Number of neurons in hidden layers: 3-50  Activation function: tanh, relu, leaky relu  Optimization algorithm: Adam, RMSprop, SGD  Learning rate: 0.01-0.00001  Batch size: 16-32  Epochs: 10-100  Regularization dropout layer with probability: 0.1-0.8  Kernel regularizer: L1, L2 |
